# High inter-home variation in nontuberculous mycobacteria from household tap waters

**DOI:** 10.64898/2026.09.06.26362288

**Authors:** Matthew J. Gebert, Jennifer E. Liggett, Caihong Vanderburgh, Jordan M. Galletta, Patrick Schwer, Teresa Muldoon, Darren A. Lytle, Sheldon V. Masters, Noah Fierer

## Abstract

Household tap waters harbor diverse microbial communities that can be highly variable in composition. While most microbes found in tap water are of limited clinical significance, nontuberculous mycobacteria (NTM), a group of bacteria commonly detected in household plumbing, pose an increasing threat to public health. Although exposure to tap water has been proposed as an important route of infection for mycobacterial pulmonary disease, it remains unclear how the amounts and types of NTM vary across homes within a given metropolitan area or even within individual homes between cold and hot tap water supplies. To address this knowledge gap, we used culture independent methods, namely 16S rRNA gene sequencing paired with quantitative PCR and mycobacterial-specific marker gene sequencing, to characterize the amounts and types of mycobacteria, including known pathogens, found in the source water, finished water, distribution system, and premise plumbing hot and cold tap waters across 16 homes in the same metropolitan area (Chicago, USA). Clinically relevant mycobacteria (*Mycobacterium avium, M. xenopi, and M. mucogenicum/phocaicum*) were frequently detected in both cold and hot tap waters, but their occurrence varied appreciably between homes receiving water from the same treatment facility. This high degree of variation in the detection of pathogenic NTM was associated with some of the measured water quality parameters, including turbidity levels, with more turbid waters being more likely to harbor pathogenic NTM, but much of the variation across tap water samples remained unexplained. The large amount of variation in the amounts and types of NTM, including those of clinically relevance, observed across households within the same water supply system highlights the need to better understand the ecology of NTM in premise plumbing to guide public health recommendations.

## Introduction

Household tap water harbors complex and diverse microbial communities, including bacteria and archaea (Thom et al. 2022; Roeselers et al. 2015; Abkar, Moghaddam, and Fowler 2024), as well as microbial eukaryotes (Perrin et al. 2019; Moreno et al. 2024) and fungi (Novak Babič and Gunde-Cimerman 2025). While most tap water-associated bacteria are innocuous, tap waters can harbor a handful of clinically significant opportunistic pathogens, including *Legionella pneumophila*, *Pseudomonas* ssp., as well as potential pulmonary pathogens within the bacterial genus *Mycobacterium*, collectively referred to as nontuberculous mycobacteria (Donohue et al. 2022; Lugli et al. 2022; Donohue et al. 2015; Dowdell et al. 2024).

Nontuberculous mycobacteria (NTM) are a group of bacteria that are nearly ubiquitous in soils, natural surface waters, household plumbing biofilms, hot tubs, tap water, and hospital water lines (Gebert et al. 2018; Lande et al. 2019; Joseph O. Falkinham 3rd 2022; Walsh et al. 2019; Modra et al. 2023). Although most mycobacterial taxa are non-pathogenic, a handful of NTM taxa can cause pulmonary disease, including *Mycobacterium avium*, *M. abscessus*, and *M. kansasii* (Daley et al. 2020). What makes these infections unique is that they are environmentally acquired, but exact mechanisms of exposure and infection from environmental reservoirs remain poorly understood, even though cases of NTM pulmonary disease and subsequent healthcare associated costs are increasing in the United States (USA) and around the world (Bents et al. 2024; Mullen et al. 2024; Dahl et al. 2022; Collier et al. 2021). Many potential sources of infection have been proposed, but residential premise plumbing (including showerheads, water heaters, and tap water) is thought to be an important route of infection (Joseph O. Falkinham 3rd 2011; Donohue et al. 2015; Gebert et al. 2018; Dowdell et al. 2024; Gebert et al. 2025).

While NTM, including those of clinical significance, have been detected in premise plumbing and tap water, previous studies have focused on either samples collected across a large geographic area (Donohue et al. 2015; Gebert et al. 2018; Donohue and Mistry 2024), or in small-scale surveys or experimental setups that may not fully capture the extent of home-to-home variability (Ji et al. 2017; Brumfield et al. 2020; Zhang et al. 2021; Rahmatika et al. 2023). Additionally, previous work has historically relied on culture dependent methods for the identification and quantification of mycobacteria (Dubrou et al. 2013; Lande et al. 2019), a strategy that can prove challenging given that NTM are notoriously difficult to cultivate *in vitro* due to their slow growth rate and the risk of overgrowth by other environmental bacteria in culture (Mercaldo et al. 2023). The amount of inter-home variability in different mycobacterial species from residential hot and cold tap waters, especially NTM of clinical significance, remains largely undetermined. This is important because the specific characteristics of household water supplies may influence the presence and associated risks of exposure to pathogenic NTM, though the determinants of differential NTM exposures remain unclear. Additionally, we have a limited understanding of how differences in water chemistry affect both the types and abundances of mycobacteria in tap water samples, an important factor to consider when designing possible interventions to reduce the risk of NTM exposures from household water supplies.

To address these knowledge gaps and assess the variation in mycobacterial exposures from tap waters within and between homes, all serviced by the same drinking water source, we collected distribution system samples along with hot and cold tap water samples across multiple time points from 16 homes within the city of Chicago, Illinois, USA. Culture independent sequencing and quantitative PCR analyses of the 16S rRNA gene paired with mycobacterial-specific marker gene sequencing (*hsp65*) were used to characterize the types and amounts of mycobacteria in water samples, including known pathogens. This study was designed to answer two overarching questions. First, how do the mycobacterial communities in these same tap water samples vary within and between homes with a focus on clinically significant strains of NTM? Second, can we identify water quality parameters that explain the observed variation in the abundances of these clinically significant NTM taxa across the homes? More generally, this study was designed to document the scale and granularity needed to better assess NTM occurrence, understand the epidemiology of NTM disease, and ultimately, better guide public health interventions moving forward.

## Methods

### Sample Collection and Processing

Tap water samples were collected from 16 residential homes in Chicago, Illinois, USA. From each home, 3 samples were collected – one from the hot tap, and two from the cold tap. All 16 homes were sampled once in April - June 2024 and again in July - September 2024, with four of the homes also sampled February - March 2024, for a total of 108 tap water samples for both chemical and microbial analyses (see Supplementary Table 1 for details).

Before collection from the cold tap, water was run with the faucet fully open for ∼30 seconds to flush. Faucet flowrates varied among collection sites, with an average of ∼4L per minute. Two 1-L bottles were pre-rinsed with cold water from the flushed tap and filled with cold water (cold stagnant). An additional 1-L of water was collected for water quality analysis. The cold-water tap was then flushed for an additional 3-5 minutes to ensure that the water sampled was from the water main. We followed an identical process as above with the flushed tap, collecting two 1-L bottles of flushed cold tap water (cold flush). For the hot tap water collection (hot stagnant), water temperature was monitored during flushing to ensure the water collected was being drawn from the storage tank. Upon the stabilization of the water temperature (between 1-8 minutes depending on the home), bottles were pre-rinsed, and two 1-L bottles were collected. Microbial collection bottles were placed on ice and transported back to the lab. Field measurements were recorded after collection (cold stagnant, cold flushed, hot stagnant): pH, temperature, free chlorine, and turbidity. Collected water was vacuum filtered within 6 hours of collection (two 1-L bottles from each collection were run through the same 0.45*μ*m polycarbonate filter), final volume was recorded, and the filters were stored at -20*°*C. Frozen filter samples were shipped on ice to the University of Colorado at Boulder for microbial processing. We used standard laboratory protocols (Campisano et al. 2017; Lipps, Braun-Howland, and Baxter 2023) to measure metal concentrations using the Agilent ICP-OES (lead, copper, cadmium, aluminum, iron, manganese, and zinc) and total phosphate and orthophosphate, using the Hach Spectrophotometer DR3900 and ThermoScientific Gallery Chemistry Analyzer, in each of the 108 tap water samples.

Water samples from the raw water intake (N=20) and finished water outlet (N=10) of the two water treatment facilities that serve all 16 homes were collected over the course of 2-weeks in September 2024. Samples were collected from continuously running taps from the two intakes (crib and shore) and an outlet at the two water treatment facilities. Two 1-L bottles were pre-rinsed with water from the tap and filled with 1-L of water from the crib intake. The identical process was performed with the shore intake and outlet, collecting two 1-L bottles of water from the tap. Collection bottles were placed on ice and transported back to the lab. Field measurements were recorded after collection (crib intake, shore intake, outlet): pH, temperature, free chlorine, and turbidity. Collected water was vacuum filtered within 6 hours of collection (two 1-L bottles from each collection were run through the same 0.45*μ*m polycarbonate filter), final volume was recorded, and the filters were stored at -20*°*C. Frozen filter samples were shipped on ice to the University of Colorado Boulder for microbial analyses. An additional 1 L of water was collected from each sample location (crib intake, shore intake, outlet) for water quality analyses as described above.

### DNA Extraction

DNA was extracted from all 138 filters (108 samples from the homes and 30 samples from the shore intake and the water purification plant outlet) using the Qiagen Powersoil Pro kit (Qiagen, Germantown, MD, USA) following manufacturer’s instructions. Collection filters were cut in half using a disposable 18G sterile needle (Air-Tite Products Co., Inc, Virginia Beach, Virginia, USA) (each needle was changed between each filter), and half the filter was used for DNA extractions. Genomic DNA was eluted in a final volume of 100*μ*L of 10 mM Tris elution buffer and stored at -20*°*C prior to downstream processing.

### 16S rRNA marker gene sequencing

To characterize the bacterial communities found in all samples we amplified the V4 hypervariable region of the 16S rRNA gene using the primer set 515F - 806R (Walters et al. 2016), with the addition of DNA extraction blanks and no-template PCR controls to check for the introduction of potential contaminants. Duplicate 25μl PCR reactions were run per sample using 12.5μl of Colorless Platinum II Hot Start Master Mix (ThermoFisher, Waltham, MA, USA), 10.5μl of PCR-grade water (Sigma-Aldrich, Burlington, MA, USA), 0.5μl 515F-barcoded primer, 0.5μl 806R primer, and 1μl of extracted genomic DNA. Thermocycler conditions were as follows: 94°C for 2 minutes, 35 cycles at 94°C for 15 seconds, 60°C for 15 seconds, 68°C for 1 minute, then 72°C for 10 minutes. Clean up and normalization were done concurrently using the Invitrogen SequalPrep Normalization Plate kit (ThermoFisher, Waltham, MA, USA) according to the manufacturer’s instructions. Five μl of cleaned and normalized amplicon from each reaction were pooled and the final library pool was quantified using the Invitrogen Qubit dsDNA HS Assay kit (ThermoFisher, Waltham, MA, USA). The library was sequenced using a 500-cycle kit on an Illumina MiSeq at the University of Colorado Boulder Center for Microbial Exploration.

### 16S rRNA gene quantitative PCR (qPCR)

To quantify the concentrations of bacteria in each of the 138 samples, we ran quantitative polymerase chain reaction (qPCR) analyses using the same 16S rRNA gene primer set (515F - 806R) as described above. We ran duplicate reactions containing 12.5 *μ*L Thermo Scientific ABsolute QPCR mix, 1.25 *μ*L forward primer (515F, 10 *μ*M), 1.25 *μ*L reverse primer (806R, *μ*M), 5*μ*L of PCR grade water, and 5 *μ*L of extracted genomic DNA, using the BIO-RAD CFX Connect Real-Time System with the following thermocycler conditions: 95*°*C for 15 minutes, followed by 94*°*C denaturation for 45 seconds, 50*°*C annealing for 1 minute, and 72*°*C elongation for 1 minute 30 seconds, repeated for 40 total cycles, followed by a 10 minute elongation step at 72*°*C. To calculate genome equivalents per sample, we generated a standard curve with serial dilutions of *Escherichia coli* genomic DNA (ATCC 700926).

Data from replicate reactions were combined and starting quantity (SQ) values from each plate were averaged. We removed cycle quantity values (Cq values) that were greater than 31 (Cq > 31) for downstream analysis, resulting in 103 tap water samples and 30 intake/outlet water samples (N=133). All qPCR data processing was done with R software (version 4.4.2, R Core Team 2024), and corresponding figures were made using the *ggplot2* package (version 3.5.1)(Wickham 2011). Final SQ values were calculated using the original volume of water that was filtered from the point-of-collection.

To calculate the abundance of mycobacterial 16S rRNA gene copies across our samples, we multiplied the total 16S rRNA gene copies per liter of water in each sample by the relative abundance of genus *Mycobacterium* found in the same sample, calculated as the proportion of 16S rRNA gene reads assigned to genus *Mycobacterium* to total 16S rRNA gene reads per sample.

### *Hsp65* gene sequencing

While the 16S rRNA sequencing and qPCR analyses allow us to quantify the amounts of the genus *Mycobacterium* in each sample, these analyses do not allow us to distinguish clinically significant species from other members of the genus not known to cause disease. To determine species level variability within the genus and to identify clinically significant NTM strains, we amplified the 65-KD heat shock protein (*hsp65*), using the Tb11-Tb12 primer set (Telenti et al. 1993). We used a “2-step” library preparation (barcodes for multiplexing are added in a second reaction, post gene amplification). In the first step, 25 μl duplicate PCR reactions were performed in reactions containing 12.5 μl of Colorless Platinum II Hot Start Master Mix (ThermoFisher, Waltham, MA, USA), 10.5 μl of PCR-grade water (Sigma-Aldrich, Burlington, MA, USA), 0.5 μl Tb11, 0.5 μl Tb12, and 1μl of extracted genomic DNA. For details regarding thermocycler conditions, see Gebert et al. (2018). Prior to the addition of the universal indices, PCR products from each first round reaction were cleaned using 0.023 μl Exo1 and 0.2275 μl SAP (New England Biolabs, Ipswich, MA) following the manufacturer’s instructions. A second PCR was run to add the universal indices to the amplicons using the same methods as described in Gebert et al. (2018). We used the identical protocol for clean-up and normalization of barcoded *hsp65* gene amplicons as described above for the 16S rRNA gene sequencing. *Hsp65* marker gene sequencing was done on a single MinION flow cell (R10.4.1, Oxford Nanopore Technologies, Oxford, UK) after library preparation with the Ligation Sequencing Amplicon V14 kit protocol (SQK-LSK114, ONT, Oxford, United Kingdom). We used super-accurate basecalling with a Min Q score of 8, generating a total of 16.22 million reads, or 10.72 Gb of sequence data from the amplicon library.

### 16S rRNA gene sequence data processing and analysis

Sequence fastq files from the MiSeq were demultiplexed with idemp (https://github.com/yhwu/idemp) and reads were trimmed to remove primer and adapter sequences using cutadapt (version 1.8.1)((Martin 2011). Data were processed using the DADA2 pipeline (version 1.32.0)(Callahan et al. 2016). Filtering was done using the quality metrics of the trimmed reads (truncLen=c(250,240), maxEE = c(2,2), truncQ=2, maxN=0). The error rates were learned, sequences were inferred (pool = TRUE, sensitive to rare taxa), and chimeric reads were removed. Taxonomy was assigned using the SILVA nr99 database (version 138.1)(Quast et al. 2013), and amplicon sequence variants (ASVs) assigned to chloroplast or mitochondria were removed from the dataset prior to downstream analyses. The final number of 16S rRNA amplicon reads after DADA2 processing and quality filtering was ∼4 million reads across 138 samples. Extraction blanks and “no-template” controls that yielded zero reads were removed from the data set. Samples with less than 1000 reads (5 water filter samples) were removed, resulting in a final dataset that consisted of 6190 ASVs detected total across 133 samples (1632 ASVs across 108 tap water samples and 5661 ASVs across 25 intake/outlet samples, with 1103 shared ASVs between the tap water and the intake/outlet samples).

### Hsp65 gene data processing and analysis

Sequence fastq files were concatenated to create one combined file for downstream processing. First, the i7 exterior Illumina adapter was trimmed off all reads using cutadapt (version 5.0)(Martin 2011) and reads were demultiplexed using dorado (version 0.9)( https://dorado-docs.readthedocs.io/en/latest/). Pre-filtering of demultiplexed reads was done using the software package chopper (version 0.8.0)(De Coster and Rademakers 2022), with read length and quality parameters based on read length distribution and quality scores (min quality (-q) = 25, min read length (-minlength) = 300, max read length (-maxlength) = 600). Sequences with ambiguous basecalls were removed from the dataset, and any extraneous primer sequences were trimmed from reads using cutadapt (version 0.8.0)((Martin 2011). The quality filtered, demultiplexed reads were then processed using the DADA2 pipeline (Callahan et al. 2016). Reads with a length less than 390 base pairs and greater than 405 base pairs were filtered from the dataset (maxEE = 2, maxLen = 405, minLen = 390). Since the Tb11-Tb12 primer set is not exclusive to genus *Mycobacterium* (i.e. it will amplify hsp65 genes from other *Actinobacteria*), we filtered the final representative sequence file using BLAST (blastn, version 2.9.0-2) (Altschul et al. 1990) against a mycobacterial *hsp65* marker gene database to include only ASVs that matched the database at 90% identity or greater. This resulted in a total of 284 mycobacterial ASVs (829701 *hsp65* reads in total) across 119 samples. Sample reads from the treatment facility intake and outlet were combined into either “intake” or “outlet” for downstream processing.

### Phylogenetic Analyses

Since many of the mycobacterial ASVs were infrequently detected and many did not have high sequence similarity to known taxa in the updated hsp65 database, we grouped ASVs into clusters based on pairwise distance, using TreeCluster (version 1.0.3)(Balaban et al. 2019), with the maximum pairwise distance allowed between two leaves no greater than 0.20 (t = 0.2). The reads in the mycobacteria-filtered representative sequence file generated from the DADA2 pipeline were aligned using MAFFT (v7.505 (2022/Apr/10)) (Katoh et al. 2002), and trimmed using trimAl (v1.4.rev15 build[2013-12-17])((Capella-Gutiérrez, Silla-Martínez, and Gabaldón 2009). We then constructed a maximum likelihood tree (ML) using RAxML (version 8.2.12, May 2018)(Stamatakis 2014), with the following command: *raxmlHPC -f a -m GTRGAMMA - p 12345 -x 12345 -# 100 -T 30.* ASV clustering resulted in the 284 mycobacterial ASVs being collapsed into 33 unique clusters, and one cluster that encompassed singleton ASVs in the dataset that did not cluster with any other group (denoted as belong to “cluster_-1”). Taxonomic identification was done by identifying those ASV clusters that included one or more known taxon in our reference database. Reads were summed together within ASV clusters and used for downstream analyses. Visualization and annotation of the resulting phylogenetic tree was done using iTOL (Interactive Tree of Life)(Letunic and Bork 2024).

### Community Level Statistical Analysis

All statistical analyses were conducted in R (R version 4.4.2, R Core Team 2024). To test whether the bacterial concentrations were higher in the intake water than the finished water, we ran a Welsh’s Two Sample t-test between the two water types, considering p<0.01 to be significant. To test for differences in bacterial concentrations (numbers of 16S rRNA gene copies per liter of water) and the water collection type (cold stagnant, cold flush, hot stagnant, N=108), a one-way analysis of variance (ANOVA) was conducted applying the Tukey’s honest significance test with a 95% family-wise confidence level (conf.level = 0.95). To determine the influence of sample type (cold stagnant, cold flushed, hot stagnant) and home site on overall bacterial community composition, we generated a dissimilarity matrix using Bray-Curtis distances followed by a permutational multivariate analysis of variance (PERMANOVA) using the *adonis2* function in the vegan package (version 2.6-8)(Oksanen et al. 2013). Principal components analysis (PCoA) was run using the phyloseq package (version 1.50.0) (McMurdie and Holmes 2013) based on pairwise Bray-Curtis dissimilarities. To determine if any correlations existed between the relative abundance of three mycobacterial taxa known to be clinically significant, and the measured water chemistry variables, we calculated the Kendall’s rank correlation coefficient for each water type between the relative abundance of an individual mycobacterial pathogen and water chemistry variables.

## Results

### General description of the bacterial communities

Although mycobacteria are the focus of this study, we first describe the results from the 16S rRNA qPCR and sequence analyses as they provide relevant baseline information on the bacterial communities in the collected water samples and context for the mycobacterial-specific results. First, as expected, the intake waters sourced from Lake Michigan had bacterial DNA concentrations that were approximately four orders of magnitude higher than concentrations observed in the finished waters collected post-treatment from the water treatment facilities (Figure 1). Likewise, bacterial DNA concentrations in household tap water samples were more similar to those observed in the finished water (Figure 1). Within homes, bacterial concentrations in both cold water collections were significantly lower than in the stagnant hot water samples (Figure 1).

**Figure 1.**
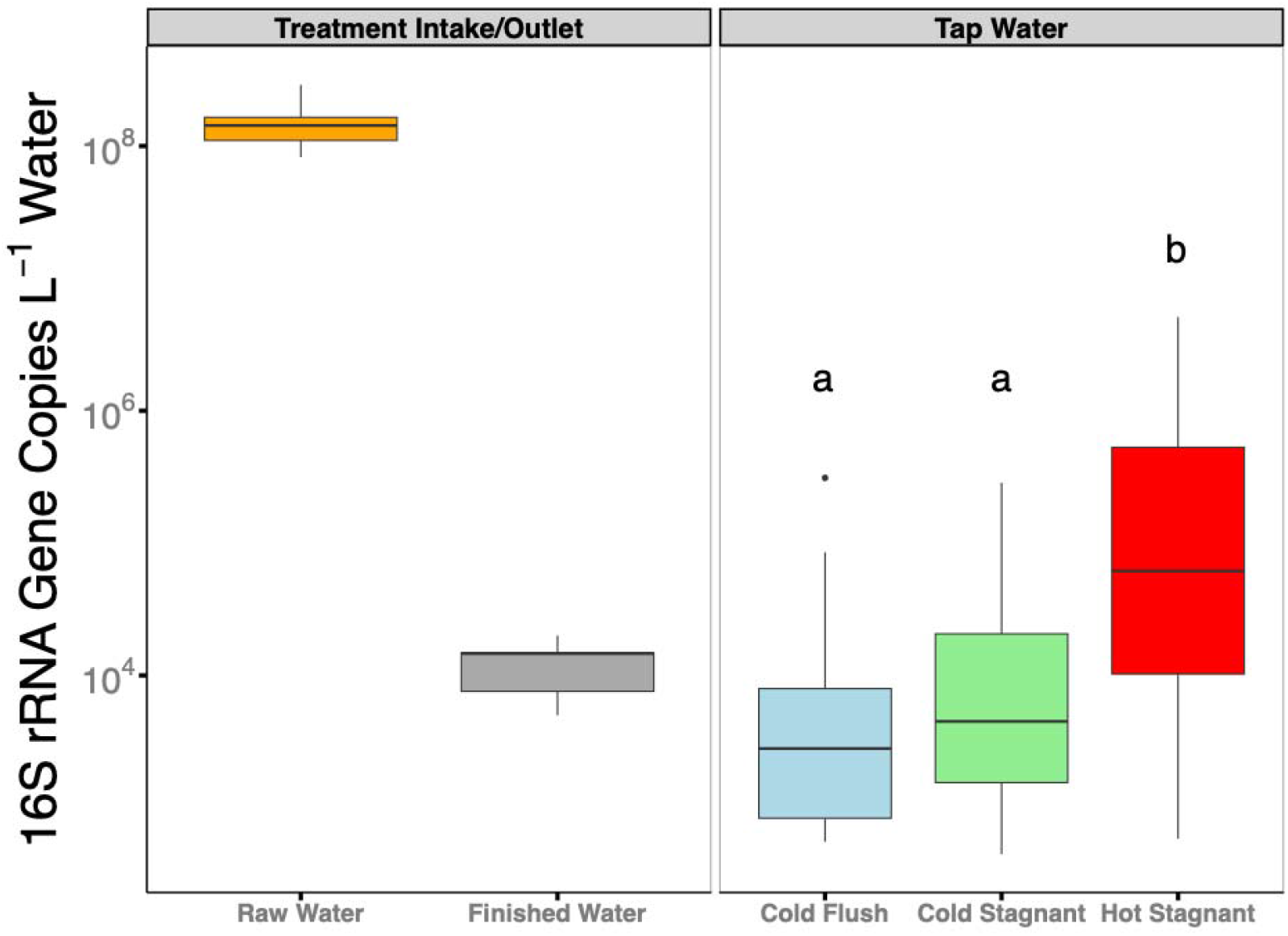
Boxplot showing the variability in bacterial DNA concentrations (16S rRNA gene copies per liter of water, determined by quantitative PCR) across tap water sample types and in the treatment intake and outlet waters. The hot tap water had a significantly higher bacterial DNA concentrations compared to the cold stagnant and cold flush samples (*ANOVA/TukeyHSD*, P = 0.003 in both cases). The two cold water tap collections were not significantly different from each other (*ANOVA/TukeyHSD*, P = 0.99). Shared letters indicate differences between water types that are not statistically significant.

We found that overall bacterial community composition in the intake waters, i.e. untreated waters, clustered distinctly from the rest of the samples (Figure 2A), while the bacterial communities in outlet waters from the treatment facility were more similar to those in the home tap waters (Figure 2B). We also found limited temporal variability in overall bacterial communities in tap waters, with different collection times per home clustering near each other in ordination space (Figure 2B), though with only 2-3 collection times per home, this study was not designed to investigate temporal variation in bacterial communities. Finally, we found that the identity of the home was a greater predictor of the tap water bacterial community composition (*PERMANOVA*, R^2^=0.35, Pr(>F) =0.001***) than within-home water source (*PERMANOVA*, R^2^=0.08, Pr(>F)=0.001***), meaning that the location of the homes in the study explained more of variation in overall bacterial community composition than the type of water collected within the home (cold stagnant, cold flush, hot stagnant).

**Figure 2.**
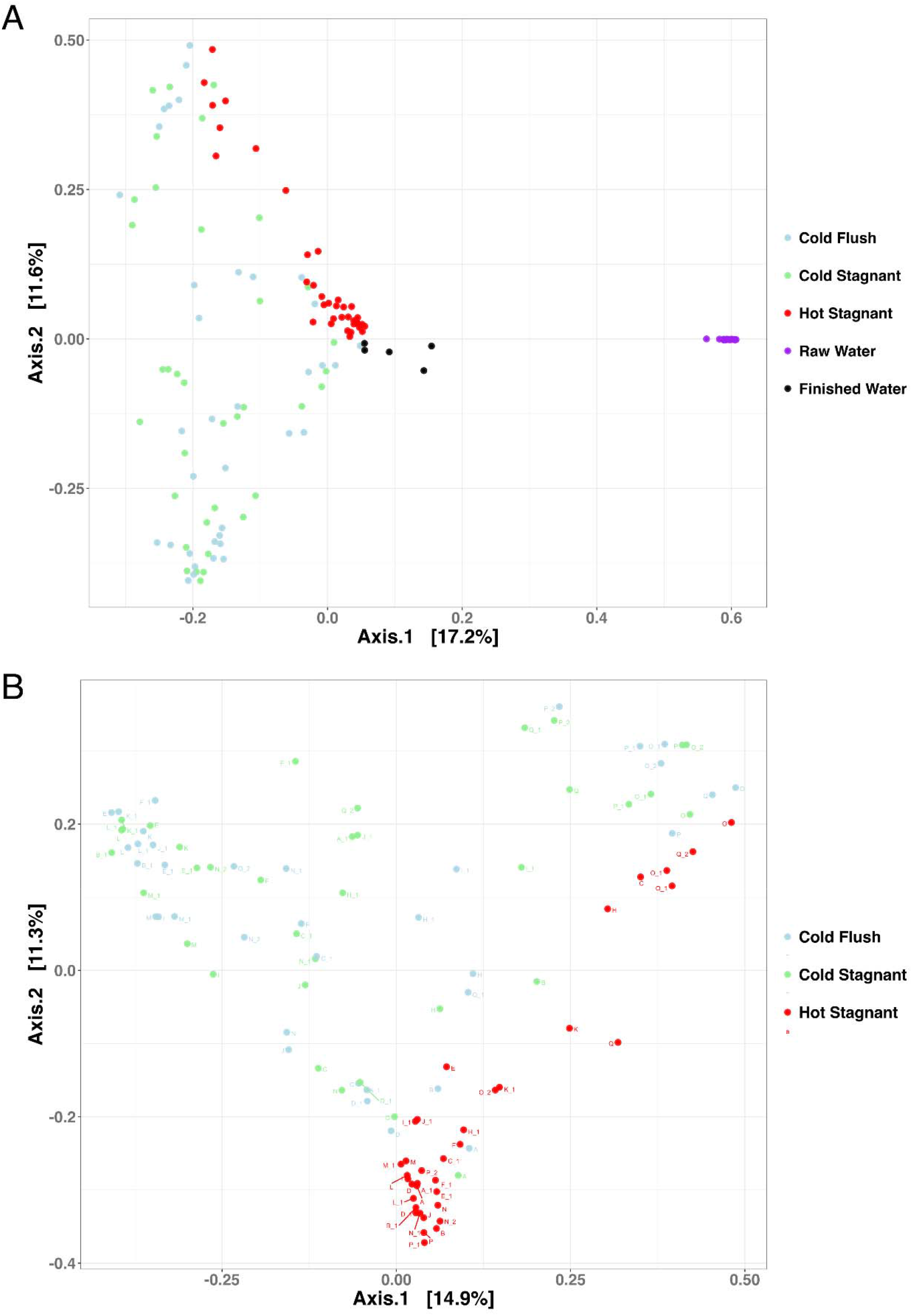
A) Principal co-ordinates analysis (PCoA), based on pairwise Bray-Curtis dissimilarities, showing the differences in overall bacterial community composition between the intake and outlet waters from the treatment facility and tap waters collected from each of the 16 homes, separated by hot and cold tap waters. The raw intake water bacterial communities (purple, untreated lake waters) cluster distinctly from the post-treatment outlet waters (black), which share more compositional similarity to the tap water samples (light blue, light green, red). B) PCoA, based on Bray-Curtis dissimilarities, showing the differences in the community composition between tap water types (cold flush, cold stagnant, and hot stagnant). Points on the plot are colored by water type and labeled alphanumerically by home and round of sample collection.

The most dominant taxonomic groups in the intake and outlet waters were the *HgcI* clade, a genus within the Phylum *Actinobacteriota,* the SAR11 clade, and genus *Cyanobium* PCC-6307 (Figure 3A). Notably, water from the intake into the treatment facility shared limited taxonomic overlap with water collected from the taps within the home. Across the three types of household tap water samples, the genus *Mycobacterium* was nearly ubiquitous in all samples and consistently the most abundant taxon (Figure 3B). The *Mycobacterium* genus was typically found to have higher relative abundances in the cold stagnant and cold flushed tap water samples than in the water samples collected from the hot water taps, but there was considerable variation across homes (Figure 3B). In addition to mycobacteria, other bacterial genera, commonly found in household plumbing systems, included *Phreatobacter*, *Sphingomonas*, *Blastomonas*, and *Bradyrhizobium* (Figure 3A).

**Figure 3.**
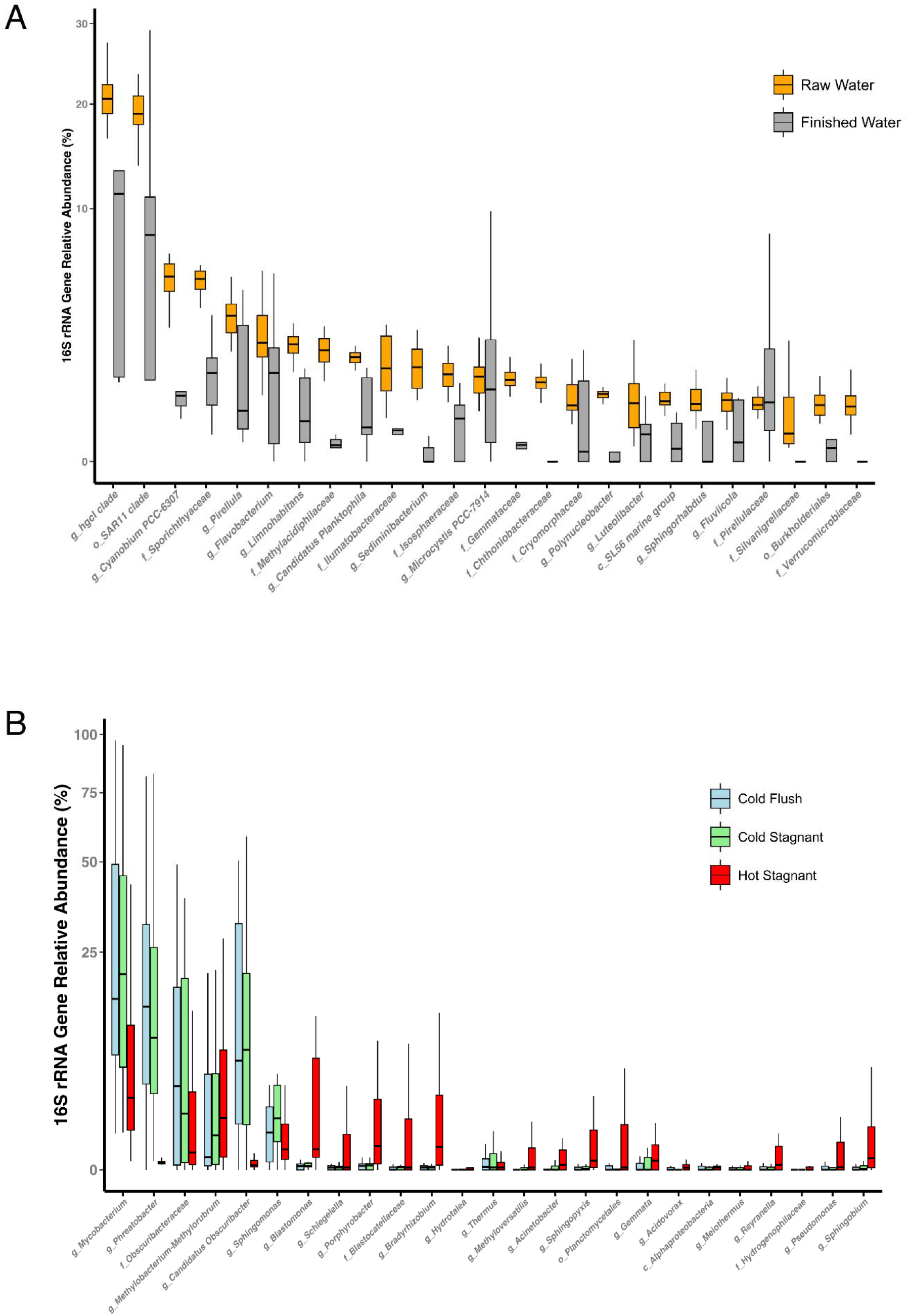
A) Boxplot showing the relative abundances, calculated as the proportion of reads matching a specific genus (‘g_’) or family (‘f_’) to the total number of 16S rRNA gene reads, for the top 25 bacterial taxa in tap water samples. Colors represent tap water sample type. B) Boxplot showing the relative abundances (calculated as in panel A) of the top 25 bacterial taxa in the treatment facility intake and outlet waters.

### Abundance of the genus Mycobacterium across source and water type

We next integrated the qPCR-derived data on total bacterial 16S rRNA gene copies with the relative abundance data for *Mycobacterium* (as determined with the same primer pairs as used for the qPCR) to determine mycobacterial concentrations in all samples. Mycobacterial DNA concentrations in the intake waters were approximately three orders of magnitude higher than in the outlet waters collected post-treatment (Figure 4A). The household tap waters had mycobacterial concentrations that were generally higher than those observed in the outlet waters, but there were no statistically significant differences across the tap water types (Figure 4A). The median mycobacterial DNA concentrations also varied substantially between homes, even across each of the tap water types within individual homes (Supplementary Figure 1).

**Figure 4.**
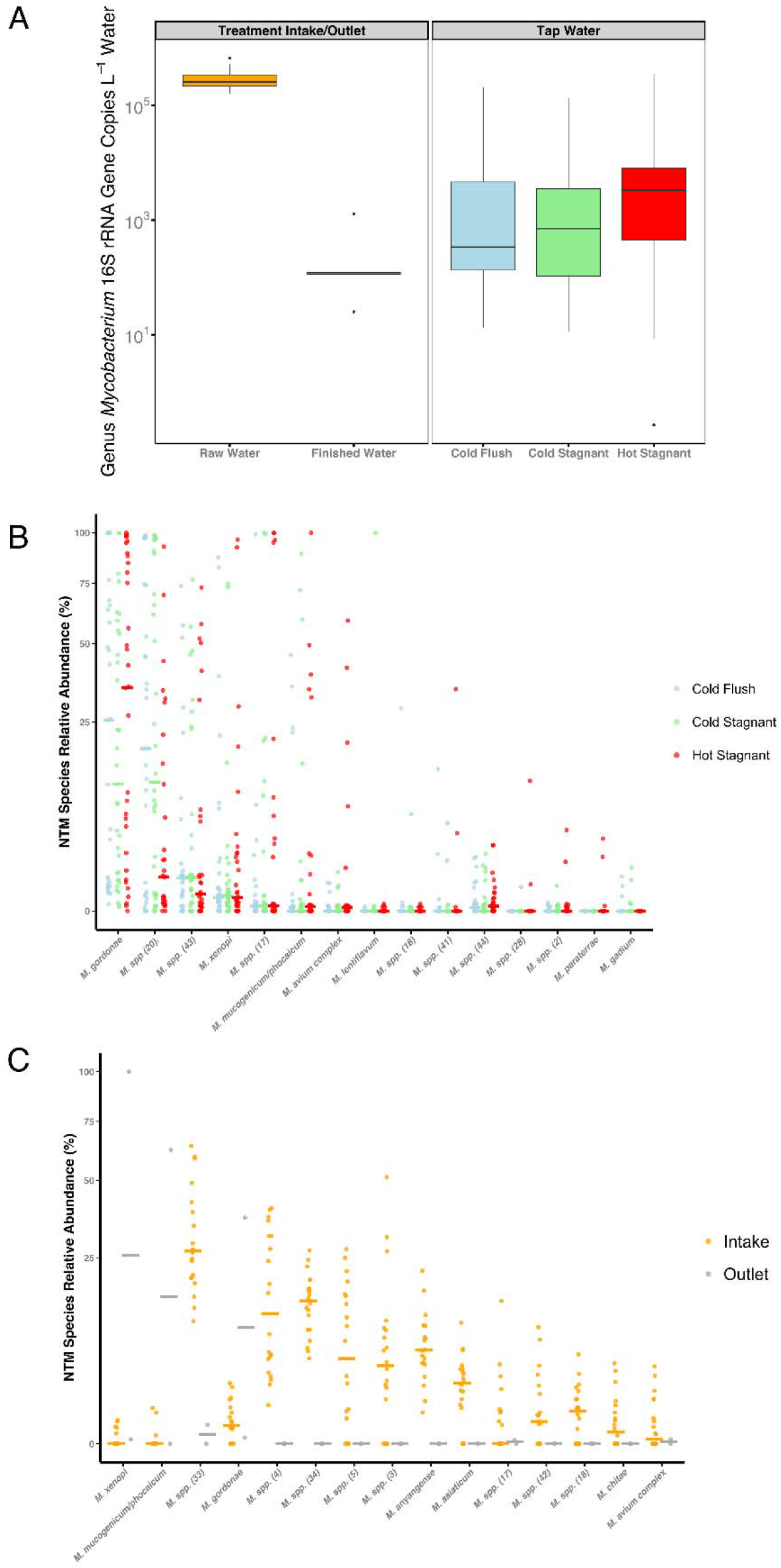
A) Boxplot showing means differences in the concentrations of the genus *Mycobacterium*, calculated as the proportion of mycobacterial reads to total 16S rRNA gene copies per liter, across tap water samples and in the treatment intake and outlet samples. Mycobacterial 16S rRNA gene copies did not vary significantly between tap water sources (*ANOVA*, p-value >0.05). B) Scatterplot showing the variation in relative abundances of the top 25 most abundant mycobacterial taxa, calculated as the proportion of species-specific *hsp65* reads assigned to each taxon to the total number of mycobacterial *hsp65* reads per sample. Points are colored by water type (light blue = cold flush, light green = cold stagnant, red = hot stagnant). C) Scatterplot showing the variation in relative abundances of the top 25 most abundant mycobacterial taxa in the treatment facility intake and outlet waters, calculated as the proportion of taxon-specific *hsp65* reads to the total number of mycobacterial *hsp65* reads per sample. Points are colored by water type (orange = intake and grey = outlet). The corresponding-colored lines in panels B and C delineate the median relative abundance of each mycobacterial taxon per water sample type.

### Mycobacterial species-level analysis via hsp65 sequencing

The genus-level analyses of mycobacterial abundances described above do not provide insight into the specific mycobacterial taxa detected in the collected water samples nor do they allow us to detect whether any of the mycobacteria are of clinical significance to humans. Thus, we paired the 16S rRNA gene analyses with *hsp65* gene sequencing to identify the types of mycobacteria, including known pathogens, in our samples. We found that *Mycobacterium gordonae* was the most dominant NTM species across the three types of tap water samples (Figure 4B). Other abundant NTM taxa included four clinically relevant groups (*Mycobacterium xenopi*, *M. mucogenicum/phocaicum*, the *M. avium* complex, and *M. lentiflavum*), but their abundances were highly variable across tap water samples (Figure 4B). The intake and outlet waters from the water treatment facility serving these homes harbored distinct mycobacterial communities from those found in the tap waters (Figure 4C) and were dominated by taxa that were distinct from any of the named species in our reference database, and therefore, could not be assigned to species, as commonly observed when conducting similar cultivation-independent analyses of the mycobacterial genus in other environmental samples (Walsh et al. 2019; Gebert et al. 2025).

### Variability in the Abundance of Clinically Relevant Mycobacteria

We next focused on three NTM taxa observed in the tap waters that are known to cause pulmonary disease (*M. xenopi*, the *M. avium* complex, and *M. mucogencium/phocaicum)*, given that exposure to these taxa could present a risk to susceptible individuals. In homes where we detected pathogenic NTM, we found a high degree of variation in both the occurrence and abundances of these clinically significant taxa between tap water types and between homes (Figure 4). *Mycobacterium xenopi* was the most ubiquitously detected of the three clinically relevant taxa, found in 50% of the homes sampled. *Mycobacterium mucogenicum/phocaicum* was the dominant pathogen in 5 of the 16 homes (31%), detected in all three water types. The *M. avium* complex was only detected in appreciable amounts in 2 of the 16 homes (12.5%) where they were found exclusively in the hot tap water samples (Figure 5).

**Figure 5.**
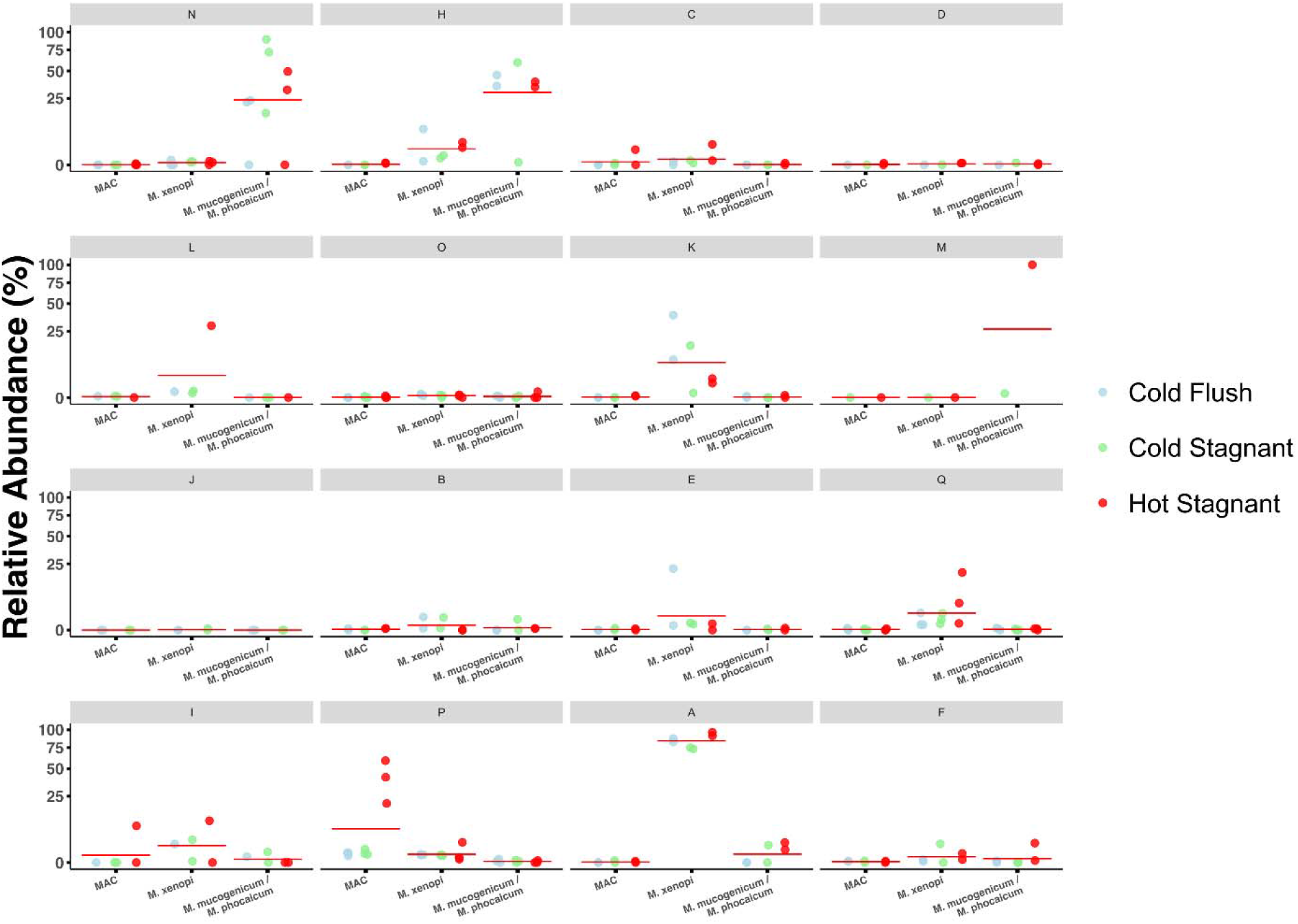
Scatterplots, separated by home, showing the variation in relative abundances, calculated as the proportion of mycobacterial *hsp65* reads assigned to each of three clinically relevant NTM taxa to the total number of mycobacterial *hsp65* reads per sample. The red lines show the median relative abundances of each clinically relevant taxon across all three sample types per home. Points are colored by tap water type.

### Environmental Predictors of Clinically Relevant NTM Abundance

Given the high variance in the detection of the three clinically relevant NTM across the homes, we next sought to determine the extent to which this variance could be explained by measured water chemistry variables. We ran correlation analyses between the relative abundances of three clinically relevant mycobacteria (*M. xenopi*, *M. avium* complex, and *M. mucogenicum/phocaicum*) and the water chemistry data (Supplementary Table 2) for each water type (cold stagnant, cold flush, hot stagnant). The abundance of *M. xenopi* was negatively correlated with free chlorine concentrations in the cold flushed tap waters (*τ* = -0.28, p-value = 0.05) and positively correlated with aluminum concentrations in the hot stagnant waters (*τ* = 0.27, p-value = 0.05). Abundances of the *M. avium* complex were positively correlated with turbidity in all three tap water types and negatively correlated with aluminum in the cold flush tap waters (*τ* = -0.3581, p-value = 0.02)(Supplementary Table 2). None of the measured water chemistry variables were associated with the relative abundance of *Mycobacterium mucogenium/phocaicum* in our dataset. For a full table of the Kendall rank correlation coefficients for the three clinically relevant strains and measured water chemistry parameters, see Supplementary Table 2.

## Discussion

Although this study largely focused on the NTM communities in household tap waters, we also investigated overall bacterial concentrations and community composition by pairing 16S rRNA gene sequencing and qPCR analyses. Not surprisingly, the intake water samples harbored had higher bacterial concentrations and distinct taxa as compared to the outlet samples collected from the water treatment plant after treatment. The intake samples were dominated by taxa we would expect to see in freshwater lake samples (e.g. the hgcI clade and SAR11 clade, (Zaremba-Niedzwiedzka et al. 2013; Henson et al. 2018; Cabello-Yeves et al. 2018; Farkas et al. 2020)), while the bacterial communities in the outlet samples were similar, but not identical, to those observed in the household tap water samples (Figure 3), highlighting that communities change in composition as water moves through the distribution network and premise plumbing (Hull et al. 2017; Bruno et al. 2022; Thom et al. 2022; Spencer-Williams et al. 2023). Across the tap water samples collected from the 16 homes, we found that bacterial concentrations were significantly higher in the hot tap water samples compared to either of the cold tap waters sample types (Figure 1), a pattern that is opposite of that observed previously (Haig et al. 2020). The hot tap water samples also had higher relative abundances of particular taxa (including *Methylobacterium*, *Blastomonas*, *Porphyrobacter*, and *Bradyrhizobium*), but similar relative abundances of the genus *Mycobacterium*, compared to the cold tap water samples (Figure 3). These results highlight that, even within an individual home, the amounts and types of bacteria found in hot tap water are distinct from those found in cold water, likely due to differences in stagnation period, temperature, and other water chemistry variables that differ between the tap water types (Supplementary Table 1).

*Mycobacterium* was the most abundant genus across both the hot and cold tap waters, consistent with previous work showing that this genus is also dominant in premise plumbing biofilm samples (Gebert et al. 2018). While we observed a sizeable decrease in mycobacterial abundances between the intake water to the treatment facility and outlet water, mycobacterial concentrations increased once they reach the both the hot and cold household tap, indicating either an accumulation in the stagnant water, or because of growth in biofilms between the treatment facility and the taps (Sousa et al. 2015; Joseph O. Falkinham 2018; Muñoz-Egea, Akir, and Esteban 2023). It is important to re-emphasize that genus-level resolution provides no insight into whether the mycobacteria detected are of clinical significance. For example, in our dataset, *Mycobacterium gordonae*, often thought to be of limited to no clinical significance (Zlojtro et al. 2015), was the most abundant species in the tap water samples, highlighting the importance of obtaining higher resolution taxonomic information to quantify exposures to clinically significant NTM.

While previous studies have sought to determine the genus level variability in mycobacteria at the household scale (Lande et al. 2019; Haig et al. 2020; Ley et al. 2020), we know of no other studies that have used cultivation independent methods to determine how the abundances and types of NTM vary across homes that share a common source. In this study, we show a high variability in clinically significant NTM, namely *M. xenopi* and *M. mucogenicum/phocaicum*, between homes, and across hot versus cold tap waters within individual homes (Figure 5). The *M. avium* complex, although rarely detected in our dataset, was found in high abundance in the hot tap water samples from 2 homes. The preferential growth of the *M. avium* complex at higher temperatures has been shown in previous studies (Schulze-Röbbecke and Buchholtz 1992; Guenette, Williams, and Falkinham 2020), yet two other clinically significant taxa, *M. xenopi* and *M. mucogenicum/phocaicum,* were detected in both the hot and cold taps, an important finding when considering potential routes of exposure in the built environment. *M. mucogenicum/phocaicum*, known primarily for causing nosocomial infections (Cooksey et al. 2008; Adékambi 2009), was detected in ∼30% of the homes in our study, a result consistent with previous work (Donohue et al. 2015). *M. xenopi*, which is often associated with higher temperatures (with optimal growth ∼40*°*C) and is known to cause pulmonary disease in humans (Marx et al. 1995; Marras et al. 2013), was detected in many of our hot and cold tap water samples.

While we observed a high degree of variation in the abundances and types of clinically significant NTM in the tap water samples collected from across the homes, this variation was not strongly associated with the water quality parameters measured at point-of-collection, so it remains largely unclear why particular homes had more of these NTM than others. However, we did identify a significant association between the presence of the *M. avium* complex in tap water samples and water turbidity. The association of *M. avium* with turbidity has been shown previously and has been hypothesized to be due to the propensity for *M. avium* to adhere to surfaces (J. O. Falkinham 3rd, Norton, and LeChevallier 2001; Joseph O. Falkinham 2018). Given the highly hydrophobic nature of NTM cell walls (Brennan and Nikaido 1995), it is likely turbidity plays an important role in both the distribution and abundance of other clinically significant NTM in premise plumbing samples, but this hypothesis needs to be tested.

The fact that we detected clinically significant NTM in tap waters highlights the important role of water treatment practices in determining NTM exposure risks. While our study does not distinguish between live and dead cells, an important distinction when considering the true risk of exposure to environmentally acquired pathogens, this study highlights the need to better understand not only the drivers of clinically relevant species in premise plumbing and tap water, but also drivers of high variability in the occurrence of these respiratory pathogens between homes all serviced by the same water source. These results also emphasize the urgent need for higher granularity in epidemiological studies of NTM occurrence from the environment. Ultimately, shifting the focus from state and county level NTM detection to household level will provide a more accurate representation of NTM exposure risks to susceptible populations and help to better inform patient and public health guidelines globally moving forward.

## Data Availability

All data produced in the present work are contained in the manuscript and supplemental material

The authors declare no conflicts of interest.

## Acknowledgements

We would like to thank all the participants who contributed water samples to this study.

## Notes

### Competing Interest Statement

The authors have declared no competing interest.

