## Supplementary Figures for "High inter-home variation in nontuberculous mycobacteria from household tap waters"

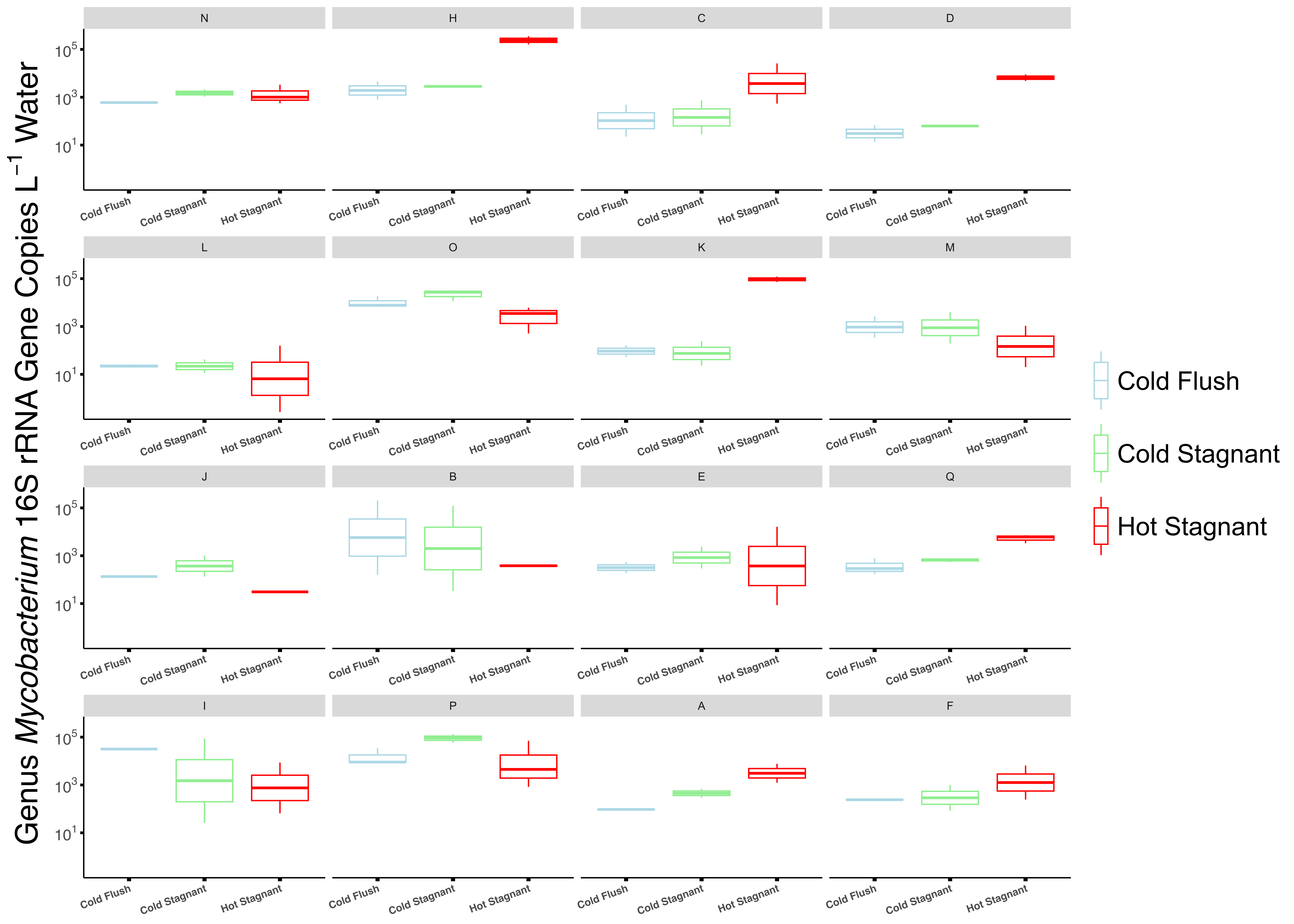

**Supplementary Figure 1.** Boxplots showing the concentrations of genus *Mycobacterium* per liter of water, as inferred from mycobacterial 16S rRNA gene copy number, across the three water sources from each home sampled.

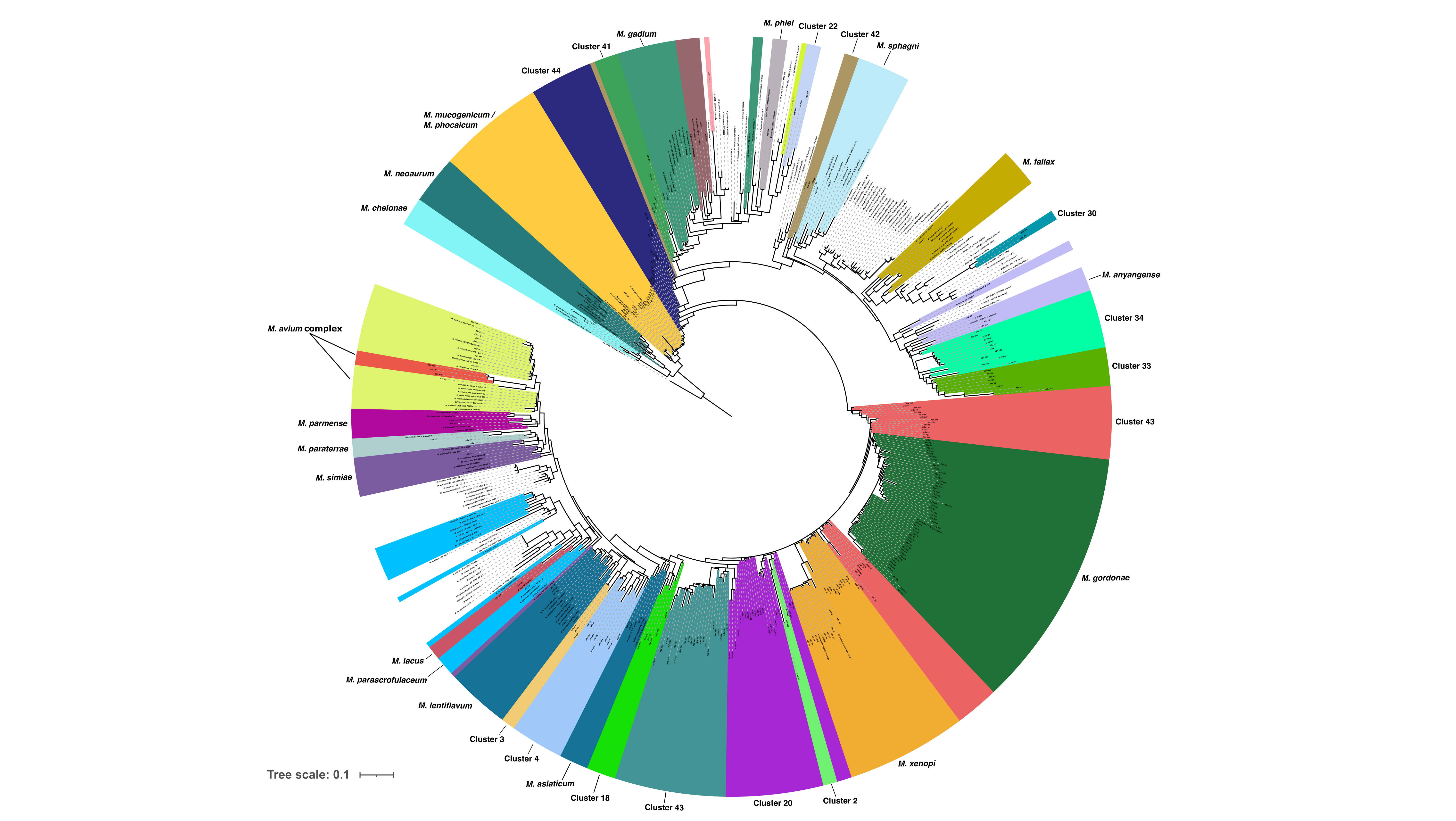

**Supplementary Figure 2.** Mycobacterial-specific *hsp65* gene maximum likelihood tree, including both ASVs and reference database sequences. Mycobacterial clusters, groups of ASVs and database sequences with <0.2 pairwise distance, are colored and labeled based on proximity to a known reference strain. The phylogenetic tree is rooted using *Nocardia farcinica* as an outgroup.
